# Elevated Pre-Treatment Systemic Immuno-Inflammatory Indices, Triple-Negative Breast Cancer, and p53 Mutation are Associated with Early-Onset Breast Cancer in Southern Nigeria

**DOI:** 10.1101/2023.11.09.23298295

**Authors:** Jude Ogechukwu Okoye, Dorcas Onyeka Samuel, Kosisochukwu Stephanie Ezidiegwu, Michael Emeka Chiemeka

## Abstract

**Background:** In West Africa, breast cancer (BC) patients have a mortality rate that is three times higher than those in North America and Northwestern Europe. This study aimed to identify high-risk patients by evaluating the pre-treatment systemic inflammatory indices, p53, and BRCA2 expressions in molecular sub-types of BC in West Africa.

**Methods:** This retrospective cohort study included 152 BC tissues, diagnosed between January 2017 and December 2022. The tissue sections were immunohistochemically stained for p53, BRCA2, hormone receptors, and human epidermal growth factor receptor 2 (HER2), scored, and analyzed accordingly. Statistical significance was set at p≤ 0.05.

**Results:** The frequency of early-onset BC (≤ 49 years) was 58.6% while the frequency of early-onset BC among patients with a family history of cancer was 76.5%. The frequency of late-stage BC was 84.9%. The frequency of luminal A and triple-negative BC (TNBC) was 1.7 times higher in early-onset BC. In comparison, the frequency of Luminal B/B-like and HER2-enriched BC was 1.9 times higher in late-onset BC (p= 0.022). The frequency of p53 and BRCA2 mutation was 1.6 times and 1.2 times higher in early-onset BC than in late-onset BC (p= 0.003 and p= 0.843, respectively). Significant differences in pre-treatment systemic inflammatory index were observed between patients with early-onset and late-onset BC, and ≤ 6 months survival and > 12 months survival (p< 0.05).

**Conclusion:** This study found a high incidence of early-onset BC, p53 mutation, and TNBC. Additionally, it suggests that pre-treatment systemic inflammatory indices can identify high-mortality-risk patients and early-onset BC.

## Introduction

Female breast cancer (BC) is the most common cancer in the world with over 2 million new cases and over 600,000 deaths annually [1]. The age-standardized incident-to-mortality ratio of BC is lower in West Africa (1.47) compared to North America and Northwestern Europe (5.12 and 4.13, respectively) [1]. This suggests that the annual fatality rate is higher in West Africa than in North America and Northwestern Europe owing to poor health infrastructure, low uptake of screening, late detection, and late-stage presentation. Triple-negative breast cancer (TNBC), also known as the basal-like breast cancer sub-type, is a highly heterogeneous disease with a poor prognosis, characterized by the absence of estrogen receptor (ER) and progesterone receptor (PR) expression and human epidermal growth factor receptor 2 (HER2) overexpression [2]. The results of a four-year study on breast cancer in Carolina show that TNBC was more common in premenopausal African-American women (39%) compared to postmenopausal African-American women (14%) and non-African-American women (16%) of all ages. The luminal-A sub-type was less common (36%) in premenopausal African-American women versus 59% and 54% in postmenopausal African-American women and non-African-American women, respectively). Their study also found that TNBC had a higher incidence of TP53 mutations and shorter survival rates [3]. DNA repair mechanisms are vital for safeguarding our cells against mutations that can lead to cancer, with deficiencies in repair pathways potentially increasing the risk of early-onset cancer. A separate study by Bauer *et al*. [4] revealed that TNBC was more prevalent among non-Hispanic black women under the age of 40. About 31-50% of hereditary BC are associated with BRCA1/2 mutations. BRCA1/2 mutations explain only about 40% of autosomal dominant inherited BCs while other genes, including TP53, explain about 10% [5]. BRCA1/2 genes encode proteins involved in repairing damaged DNA [6]. BRCA2 expression significantly correlates with grade III, suggesting that overexpression of BRCA2 may influence the aggressiveness of breast tumours. In a mouse model, the inactivation of BRCA2 and p53 combine to mediate mammary tumorigenesis, and p53 pathway inactivation precedes BRCA2-associated breast tumour formation. Immunohistochemical loss of the BRCA gene is indicative of BRCA dysfunction with high sensitivity and specificity of 80-90% [6,7]. Inflammatory cells, when chronically activated, can contribute to DNA damage and inflammation-driven cancer. The interplay between these factors is particularly relevant in the context of early-onset cancer, where genetic predispositions may converge with DNA repair processes and inflammatory responses to promote tumorigenesis at a younger age which in the long run determines patients’ survival rates [8]. Research has shown that neutrophils and monocytes can contribute to tumour growth and impede immune surveillance, whereas lymphocytes play a vital role in preventing immune evasion. A correlation has been observed between elevated counts of neutrophils and monocytes and reduced counts of lymphocytes with the onset and advancement of tumours [9]. Quantifying the inflammatory response can be used as an affordable prognostic tool to assess disease outcomes. This study is the first to compare the systemic inflammatory indices, p53, and BRCA2 expressions, hormone receptors and HER2 in early-onset and late-onset breast cancer in Southern Nigeria.

## Materials and Methods

### Study Population

This retrospective study included 152 cases of BC diagnosed from January 2017 to December 2022 at the Department of Gynaecology, Nnamdi Azikiwe University Teaching Hospital and private clinics in Nnewi and Onitsha, Nigeria. Patients with secondary breast cancer were excluded from the study. Some patients received Cyclophosphamide, Doxorubicin/Adriamycin, and 5-fluorouracil as neoadjuvant chemotherapy. Some patients received second-line chemotherapy (Paclitaxel/Docetaxel and carboplatin/capecitabine). The patient’s medical records were accessed for socio-clinical demographics such as age, gender, comorbidities, and time of presentation. All analyses were performed by the ethical standards laid down in the Declaration of Helsinki. Patients were grouped into early-onset BC and late-onset BC.

### Sample collection and handling

Each patient provided two samples of 5 ml of venous whole blood, which were collected and discharged into EDTA containers - one a week before the first chemotherapy and the other a week before discharge. The whole blood samples were analyzed using a Haemo-autoanalyzer to obtain full blood counts. After ultrasound investigations, biopsy, and surgery, the resected tissues were sent to the Department of Morbid Anatomy and Forensic Medicine for histological investigation. At least two pathologists evaluated the tissues for evidence of malignancy and the disease was staged based on the American Cancer Society guidelines [10]. The following parameters were calculated for the subgroups: total white cell count (TWBC 10^9/L), neutrophil-to-lymphocyte ratio (NLR), platelet-to-lymphocyte ratio (PLR), platelets-neutrophils to lymphocytes ratio (PNLR; [Platelet count x Neutrophil count]/Lymphocyte count), and neutrophils-to-lymphocytes-platelets ratio (NLPR; [Neutrophil count x 100]/Lymphocyte count x platelet count).

### Immunohistochemical technique

Cases of BC were subclassified into 3 groups based on keratinization, invasiveness, and extent of differentiation. The presence of ER, PR, HER2, BRCA2, and p53 proteins in the tissue sections was determined by immunohistochemical technique as described by Buchwalow and Bocker [11]. Sections from confirmed positive and negative cases were used as controls. Sections of 3 microns from formalin-fixed paraffin-embedded tissue samples were mounted on charged slides and air-dried for 2 hours at 60°C. Sections were deparaffinized in three changes of xylene and hydrated through grades of alcohol. Thereafter, tissue sections were subjected to heat epitope retrieval using citrate buffer at pH 6.0 at 95°C. The retrieval solution was heated in a water bath to 65°C before the slides were introduced and heated to 95°C. The sections and buffer were further heated at 95°C for 20 mins. Sections were then allowed to cool at room temperature for 20 mins and adequately washed using phosphate-buffered saline (PBS) at pH 7.4. This was followed by placing the tissue sections in a peroxidase blocker, allowing them to stand for 5 minutes, and subsequently washing them using PBS. The circumferences of tissue sections on slides were marked round with a grease pencil and subsequently covered with primary antibodies (diluted with immunodetection protein blocker/antibody diluent), allowed to stand for 60 mins, and adequately washed using PBS. Tissue sections further were covered with a biotin link, allowed to stand for 10 minutes, and washed using PBS. Subsequently, tissue sections were covered with a horseradish peroxidase label, incubated for 10 mins, and washed with deionized water. Tissue sections were covered with DAB substrate-chromogen solution (one drop of DAB chromogen in one ml of immunodetection DAB buffer), allowed to stand for 5 mins, and rinsed with deionized water. The sections were counterstained with haematoxylin for 2 mins, rinsed in PBS and water, dehydrated, dealcoholized in xylene, and permanently mounted using coverslips. Photomicrographs were taken for documentation. Immunoglobin G1-based primary antibodies included ER, PR, HER2, BRCA2, and p53. Positive staining (brown) in the tissues was scored using a scale of 0, +1, +2, and +3, based on intensity. Scores 0 and +1 were considered negative while scores +2 and +3 were considered positive [12].

### Statistical analysis

Chi-square/Fisher was used to determine the association between the socio-clinical demographics of patients who were ≤ 49 years old and those > 49 years of age. Pearson’s correlation was used to determine the relationship between the variables (NLR, PLR, PNLR, NLPR, and LMR) before treatment. A T-test was used to compare data of 1. patients aged ≤ 49 years old (early onset) and those > 49 years (late-onset), ANOVA was used to compare patients’ data based on duration in care (DIC); ≤ 6 months, 7 -12 months, and > 12 months.

## Results

The mean and median ages of the patients were 48.93 ± 13.89 years and 45.10 years, respectively. According to the results, the highest number of BC cases were diagnosed in 2019, specifically within the age group of 40 to 49 years (as illustrated in Figure 1). The survival rates of patients before the COVID-19 pandemic declaration (2017-2019) and after (2020-2021) were 299.0 ± 31.8 days and 205.5 ± 23.8 days, respectively (p= 0.022). The frequency of early-onset cases was lower in the former (56.2%) than in the latter (63.9%), but this difference was not statistically significant (p= 0.408).

**Figure 1:**
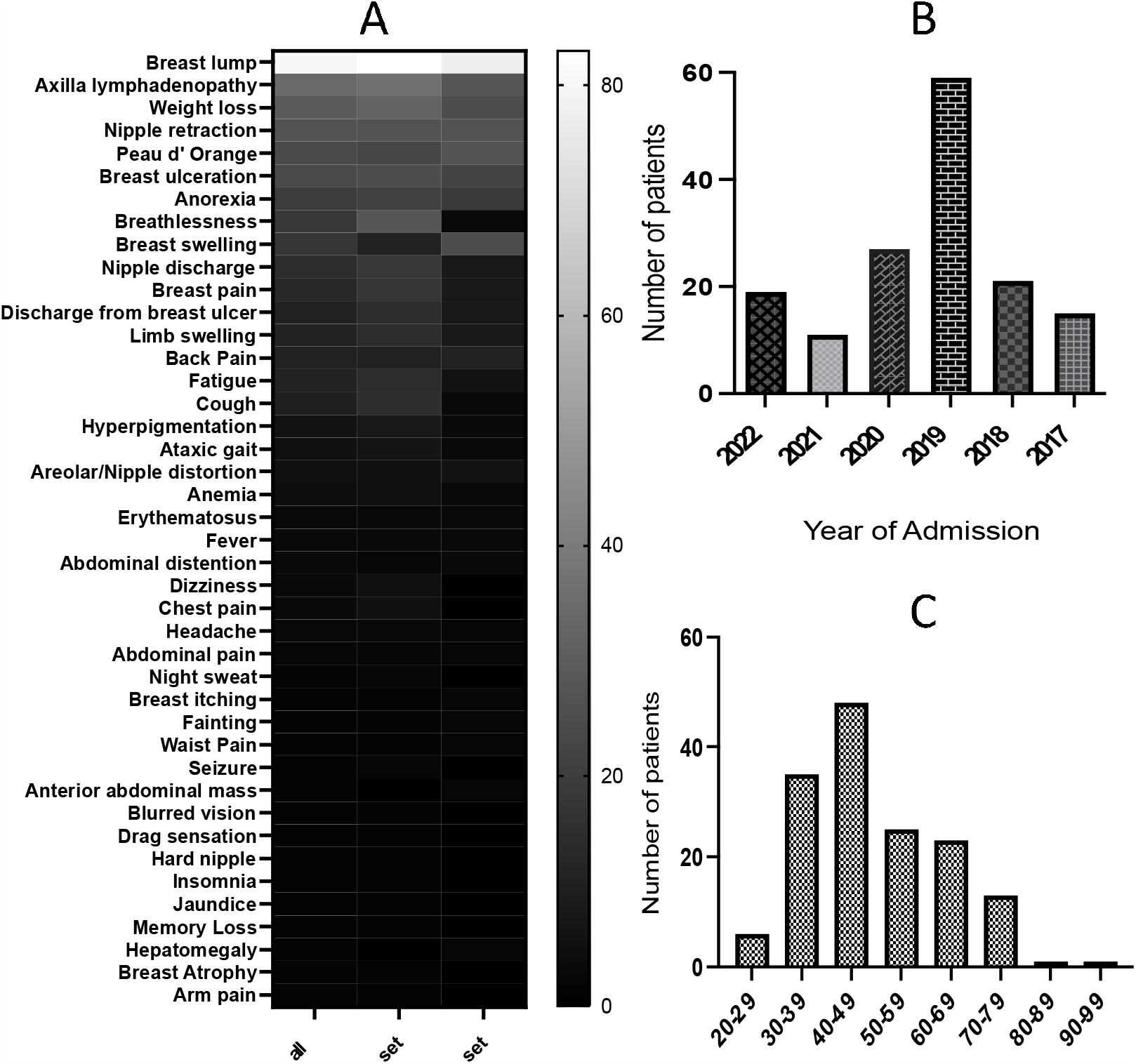
Features, year of diagnosis and age distribution of patients diagnosed with BC. Figure 1A shows that breast lump and axilla lymphadenopathy were the most frequent signs present by the patients (80.9% and 33.6%, respectively). Figure 1B indicates that the year 2019 had the highest frequency of BC diagnoses, followed by 2020. Figure 1C revealed that the occurrence of BC was most prevalent in the age group of 40 to 49 years. Additionally, early-onset BC (20 to 49 years) was found to be more frequent than late-onset BC (50 to 99 years)

### Socio-demographic features

The age range was 23 to 96 years. Results in Table 1 show that the unemployment rate was 2.0 times lower among patients aged 49 years old and below (Group A) compared with patients who were over 49 years old (Group B) at p> 0.05. The frequency of secondary and tertiary education was 1.7 times higher in Group A compared with Group B (p< 0.05). The uptake of a complete course of first-line and second-line chemotherapy was lower among unemployed patients (13.6% and 13.6%) compared with their employed counterparts (42.4% and 25.6%) at p= 0.016 and p= 0.4100, respectively. The alcohol consumption rate was slightly higher in Group A compared to Group B (p>0.05), while tobacco use was 10.1 times more prevalent in Group B than in Group A (p<0.05). The frequency of hypertension and diabetes was higher in Group B than in Group A (1.1 and 3.7 times, respectively; p>0.05 and <0.05). In contrast, a history of pre-hypertension was 1.4 times higher in Group A compared with Group B (p> 0.05). The consumption of herbal therapy was slightly higher in Group B than in Group A (p>0.05). The history of miscarriage or fetal death was 1.7 times higher in Group A compared to Group B (p> 0.05). The history of oral conceptive uptake was 1.8 times higher in Group A compared to Group B (p> 0.05). The frequency of obesity and early menarche (between ages 12 and 13) were 1.2 and 1.6 times higher, respectively, in Group B than in Group A (p < 0.05). Additionally, the frequency of early-age pregnancy (between ages 10 and 19) was 2.0 times higher in Group B compared to Group A (p < 0.05). The occurrence of multiple births was 1.3 times greater in Group B compared to Group A (p< 0.05). Moreover, the frequency of breastfeeding for more than 6 months was 1.2 times higher in Group A than in Group B (p< 0.05).

**Table 1:** Socio-demographic characteristics of patients diagnosed with breast cancer.

| Variables | No. (%) | ≤ 49 years | > 49 years | X<sup>2</sup> | p-value |
| --- | --- | --- | --- | --- | --- |
|  | N= 152 | n= 89 (%) | n= 63 (%) |  |  |
| **Employment status:** |  |  |  | 5.232 | 0.073 |
| Civil servant | 33 (21.7) | 18 (20.2) | 15 (23.8) |  |  |
| Self employed | 96 (63.2) | 62 (69.7) | 34 (54.0) |  |  |
| Unemployed | 23 (15.1) | 9 (10.1) | 14 (22.2) |  |  |
| **Level of Education:** |  |  |  | 28.21 | <0.001* |
| No formal education | 4 (2.6) | 0 (0.0) | 4 (6.3) |  |  |
| Primary education | 35 (23.0) | 9 (10.1) | 26 (41.3) |  |  |
| Secondary education | 63 (41.4) | 45 (50.6) | 18 (28.6) |  |  |
| Tertiary education | 50 (32.9) | 35 (39.3) | 15 (23.8) |  |  |
| **Alcohol consumption:** |  |  |  | 0.186 | 0.666 |
| No | 118 (77.6) | 68 (76.4) | 50 (79.4) |  |  |
| Yes | 34 (22.4) | 21 (23.6) | 13 (20.6) |  |  |
| **Tobacco Use:** |  |  |  | 7.380 | 0.007* |
| No | 144 (94.7) | 88 (98.9) | 56 (88.9) |  |  |
| Yes | 8 (5.3) | 1 (1.1) | 7 (11.1) |  |  |
| **History of Hypertension:** |  |  |  | 0.885 | 0.643 |
| None | 65 (42.8) | 38 (42.7) | 27 (42.9) |  |  |
| Pre-HTN | 24 (15.8) | 16 (18.0) | 8 (12.7) |  |  |
| HTN | 63 (41.4) | 35 (39.3) | 28 (44.4) |  |  |
| **Diabetes Mellitus** |  |  |  | 4.781 | 0.029* |
| No | 141 (92.8) | 86 (96.6) | 55 (87.3) |  |  |
| Yes | 11 (7.2) | 3 (3.4) | 8 (12.7) |  |  |
| **History of Herbal therapy:** |  |  |  | 0.0281 | 0.867 |
| No | 124 (81.6) | 73 (82.0) | 51 (81.0) |  |  |
| Yes | 28 (18.4) | 16 (18.0) | 12 (19.0) |  |  |
| **History of Miscarriage/fetal death** |  |  |  | 0.982 | 0.322 |
| No | 141 (92.8) | 81 (91.0) | 60 (95.2) |  |  |
| Yes | 11 (7.2) | 8 (8.0) | 3 (4.8) |  |  |
| **History of Oral contraceptives** |  |  |  | 0.501 | 0.479 |
| No | 145 (95.4) | 84 (94.4) | 61 (96.8) |  |  |
| Yes | 7 (4.6) | 5 (5.6) | 2 (3.2) |  |  |
| **Body Mass Index (BMI)** |  |  |  | 8.101 | 0.004* |
| ≤ 24.9 | 53 (34.9) | 35 (39.3) | 18 (28.6) |  |  |
| >24.9 | 99 (65.1) | 54 (60.7) | 45 (71.4) |  |  |
| **Menarche:** |  |  |  | 8.068 | 0.018* |
| 12-13 years | 60 (39.5) | 28 (31.5) | 32 (50.8) |  |  |
| 14-15 years | 58 (38.2) | 35 (39.3) | 23 (36.5) |  |  |
| ≥16 years | 34 (22.4) | 26 (29.2) | 8 (12.7) |  |  |
| **Age at first confinement** |  |  |  | 9.316 | 0.010* |
| 10 – 19 | 18 (11.8) | 8 (8.0) | 10 (15.9) |  |  |
| 20 – 29 | 90 (59.2) | 47 (52.8) | 43 (68.2) |  |  |
| ≥ 30 | 42 (27.6) | 34 (38.2) | 10 (15.9) |  |  |
| **Parity:** |  |  |  | 22.25 | <0.001 |
| 0 | 23 (15.1) | 17 (19.1) | 6 (9.5) |  |  |
| ≤ 2 | 24 (15.8) | 18 (20.2) | 6 (9.5) |  |  |
| 3 – 4 | 49 (32.2) | 35 (39.3) | 14 (22.2) |  |  |
| > 4 | 56 (36.8) | 19 (21.3) | 37 (58.7) |  |  |
| **Average breast feeding** |  |  |  | 6.449 | 0.040* |
| ≤ 6 months | 49 (32.2) | 25 (28.1) | 24 (38.1) |  |  |
| 7 – 12 months | 55 (36.2) | 39 (43.8) | 16 (25.4) |  |  |
| > 12 months | 48 (31.6) | 25 (28.1) | 23 (36.5) |  |  |

TSMP: Time of symptom manifestation to presentation. Descriptive analysis and Chi-square/Fisher's exact test. \*Significance was set at p < 0.05.

### Clinical Characteristics

Table 2 shows that early-onset breast cancer had a 1.4 times higher incidence rate than late-onset BC. The table also demonstrates that late-stage BC was 5.6 times more common than early-stage BC. The frequency of family history of (breast and prostate) cancer was 1.7 times higher in Group A (patients 49 years or under) compared with Group B (patients older than 49 years) at p> 0.05. In other words, 76.5% of patients with a family history of any cancer were of early-onset type. Among the cohort, the frequency of late presentation (symptom-wise) was high although Group B exhibited a higher frequency of early presentation at the clinic when compared to Group A (p> 0.05). Additionally, it was found that the frequency of right breast cancer was higher than that of left breast cancer. Interestingly, the frequency of bilateral breast cancer was 5.6 times higher in Group A than in Group B (p= 0.080). The frequency of poorly differentiated and high-score BC (6 to 9) of Group A were 1.2 and 1.1 times higher than in Group B, respectively (p> 0.05). The frequency of invasive BC was 1.1 times higher in Group B than in Group A (p> 0.05). The study also found that the frequency of Luminal A and triple-negative breast cancer was 1.7 times higher in Group A compared to Group B. On the other hand, the frequency of Luminal B/B-like and HER2-enriched cancer was 1.9 times higher in Group B than in Group A (p< 0.05). The frequency of p53 and BRCA2 mutations in Group A was 1.6 times and 1.2 times higher than in Group B, respectively (p<0.05 and p>0.05). The average age of patients diagnosed with TNBC was found to be 48.92 ± 13.61 years, which is lower than that of patients diagnosed with Luminal B-like cancer (53.0 ± 11.01 years). The prevalence of p53/BRCA2 loss in TNBCs and Luminal B-like cancer was 68.7%/46.9% and 81.0%/33.3%, respectively indicating that p53 loss precedes BRCA2 loss. The mutation of both p53 and BRCA2 were higher in TNBCs (15.6%) than in Luminal B-like cancer (11.9%). The frequency of stage 4 BC was 1.2 times higher in Group A compared to Group B (p= 0.497). Only 47% of the 117 patients who received chemotherapy completed six courses. Group A had a 1.2 times higher number of chemotherapy-naïve patients than Group B (p> 0.05). Group A and Group B had similar in-hospital death rates of 19.1% and 18.0%, respectively.

**Table 2:**
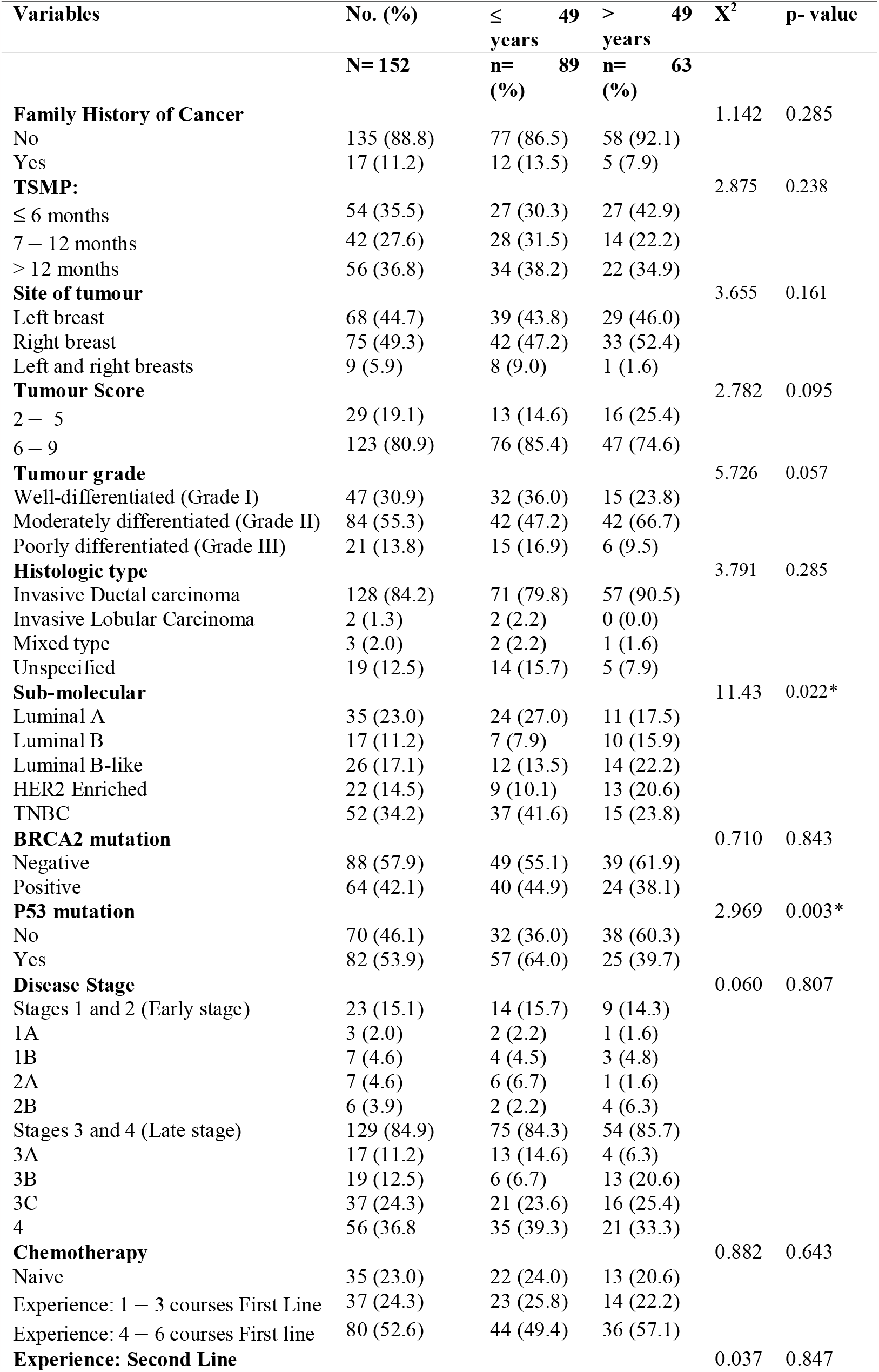
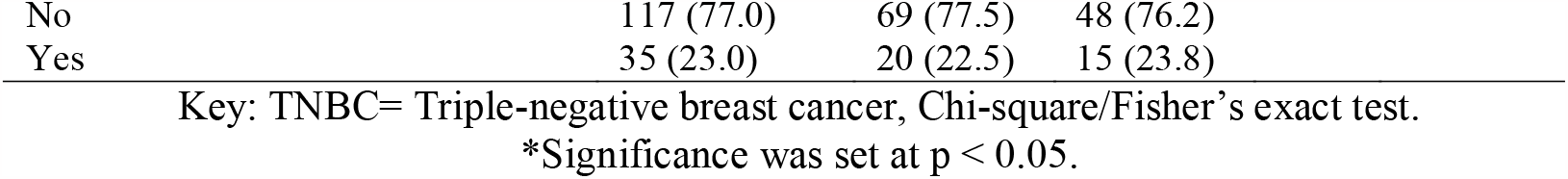
Clinical characteristics of patients diagnosed with breast cancer.

| Variables | No. (%) | ≤ 49 years | > 49 years | X<sup>2</sup> | p-value |
| --- | --- | --- | --- | --- | --- |
|  | N= 152 | n= 89 (%) | n= 63 (%) |  |  |
| **Family History of Cancer** |  |  |  | 1.142 | 0.285 |
| No | 135 (88.8) | 77 (86.5) | 58 (92.1) |  |  |
| Yes | 17 (11.2) | 12 (13.5) | 5 (7.9) |  |  |
| **TSMP:** |  |  |  | 2.875 | 0.238 |
| ≤ 6 months | 54 (35.5) | 27 (30.3) | 27 (42.9) |  |  |
| 7 – 12 months | 42 (27.6) | 28 (31.5) | 14 (22.2) |  |  |
| > 12 months | 56 (36.8) | 34 (38.2) | 22 (34.9) |  |  |
| **Site of tumour** |  |  |  | 3.655 | 0.161 |
| Left breast | 68 (44.7) | 39 (43.8) | 29 (46.0) |  |  |
| Right breast | 75 (49.3) | 42 (47.2) | 33 (52.4) |  |  |
| Left and right breasts | 9 (5.9) | 8 (9.0) | 1 (1.6) |  |  |
| **Tumour Score** |  |  |  | 2.782 | 0.095 |
| 2 – 5 | 29 (19.1) | 13 (14.6) | 16 (25.4) |  |  |
| 6 – 9 | 123 (80.9) | 76 (85.4) | 47 (74.6) |  |  |
| **Tumour grade** |  |  |  | 5.726 | 0.057 |
| Well-differentiated (Grade I) | 47 (30.9) | 32 (36.0) | 15 (23.8) |  |  |
| Moderately differentiated (Grade II) | 84 (55.3) | 42 (47.2) | 42 (66.7) |  |  |
| Poorly differentiated (Grade III) | 21 (13.8) | 15 (16.9) | 6 (9.5) |  |  |
| **Histologic type** |  |  |  | 3.791 | 0.285 |
| Invasive Ductal carcinoma | 128 (84.2) | 71 (79.8) | 57 (90.5) |  |  |
| Invasive Lobular Carcinoma | 2 (1.3) | 2 (2.2) | 0 (0.0) |  |  |
| Mixed type | 3 (2.0) | 2 (2.2) | 1 (1.6) |  |  |
| Unspecified | 19 (12.5) | 14 (15.7) | 5 (7.9) |  |  |
| **Sub-molecular** |  |  |  | 11.43 | 0.022* |
| Luminal A | 35 (23.0) | 24 (27.0) | 11 (17.5) |  |  |
| Luminal B | 17 (11.2) | 7 (7.9) | 10 (15.9) |  |  |
| Luminal B-like | 26 (17.1) | 12 (13.5) | 14 (22.2) |  |  |
| HER2 Enriched | 22 (14.5) | 9 (10.1) | 13 (20.6) |  |  |
| TNBC | 52 (34.2) | 37 (41.6) | 15 (23.8) |  |  |
| **BRCA2 mutation** |  |  |  | 0.710 | 0.843 |
| Negative | 88 (57.9) | 49 (55.1) | 39 (61.9) |  |  |
| Positive | 64 (42.1) | 40 (44.9) | 24 (38.1) |  |  |
| **P53 mutation** |  |  |  | 2.969 | 0.003* |
| No | 70 (46.1) | 32 (36.0) | 38 (60.3) |  |  |
| Yes | 82 (53.9) | 57 (64.0) | 25 (39.7) |  |  |
| **Disease Stage** |  |  |  | 0.060 | 0.807 |
| Stages 1 and 2 (Early stage) | 23 (15.1) | 14 (15.7) | 9 (14.3) |  |  |
| 1A | 3 (2.0) | 2 (2.2) | 1 (1.6) |  |  |
| 1B | 7 (4.6) | 4 (4.5) | 3 (4.8) |  |  |
| 2A | 7 (4.6) | 6 (6.7) | 1 (1.6) |  |  |
| 2B | 6 (3.9) | 2 (2.2) | 4 (6.3) |  |  |
| Stages 3 and 4 (Late stage) | 129 (84.9) | 75 (84.3) | 54 (85.7) |  |  |
| 3A | 17 (11.2) | 13 (14.6) | 4 (6.3) |  |  |
| 3B | 19 (12.5) | 6 (6.7) | 13 (20.6) |  |  |
| 3C | 37 (24.3) | 21 (23.6) | 16 (25.4) |  |  |
| 4 | 56 (36.8) | 35 (39.3) | 21 (33.3) |  |  |
| **Chemotherapy** |  |  |  | 0.882 | 0.643 |
| Naive | 35 (23.0) | 22 (24.0) | 13 (20.6) |  |  |
| Experience: 1 – 3 courses First Line | 37 (24.3) | 23 (25.8) | 14 (22.2) |  |  |
| Experience: 4 – 6 courses First line | 80 (52.6) | 44 (49.4) | 36 (57.1) |  |  |
| **Experience: Second Line** |  |  |  | 0.037 | 0.847 |

|  |  |  |  |
| --- | --- | --- | --- |
| No | 117 (77.0) | 69 (77.5) | 48 (76.2) |
| Yes | 35 (23.0) | 20 (22.5) | 15 (23.8) |

Key: TNBC= Triple-negative breast cancer, Chi-square/Fisher's exact test.
\*Significance was set at $p < 0.05$ .

Approximately 49% and 62% of patients in Groups A and B, respectively, received care for over 6 months. Elevated platelet-related systemic immune-inflammatory indices were observed among patients with early-onset BC compared to patients with late-onset BC (Figure 2).

**Figure 2:**
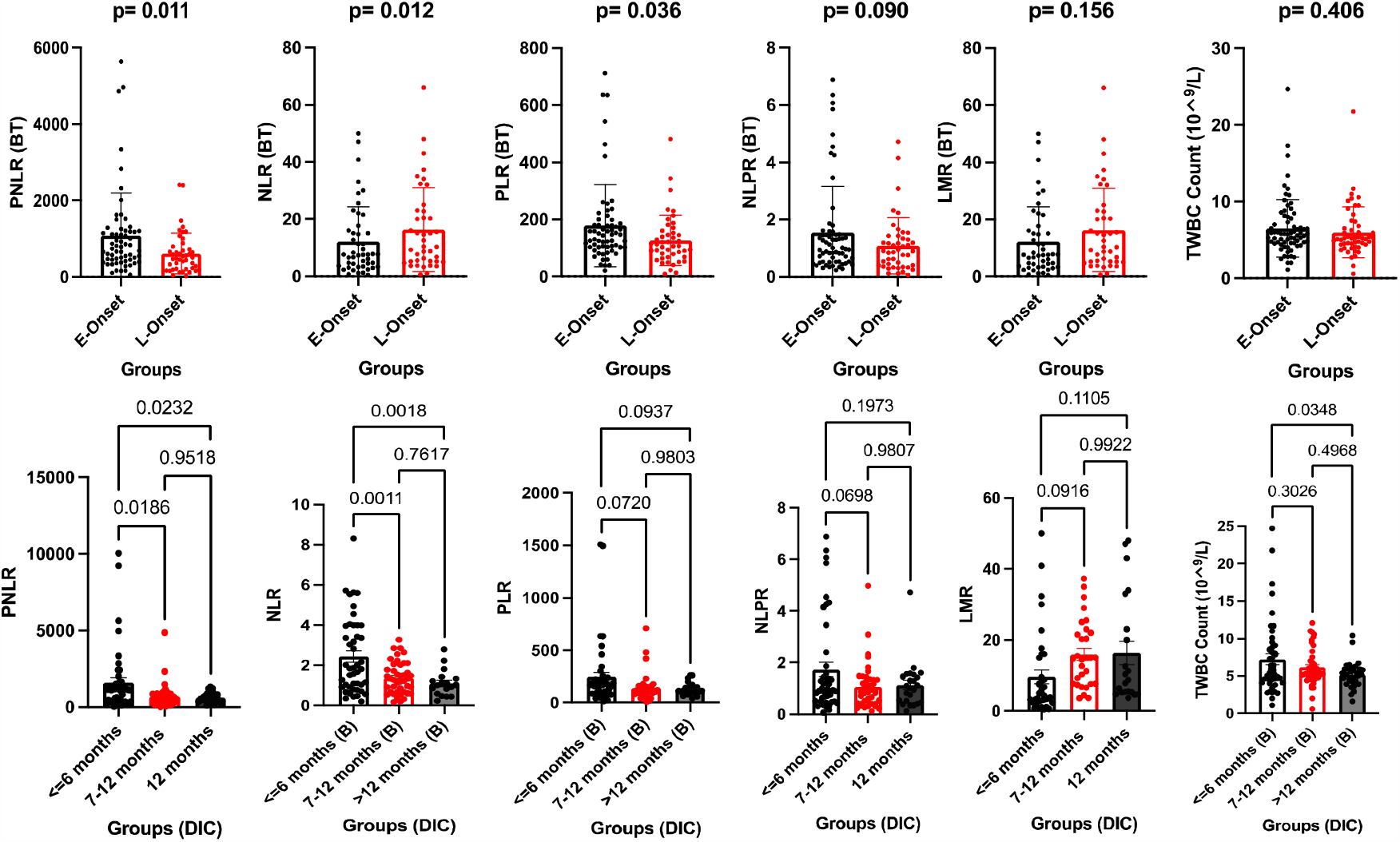
Comparison of pre-treatment white cell count in early-onset and late-onset BC. Figure 2 shows significantly lower pre-treatment (BT) PNLR and PLR, and insignificant lower pre-treatment NLPR and TWBC among patients with late-onset (L-onset) BC compared to their counterparts with early-onset (E-onset) BC at p< 0.05, < 0.05, > 0.05 and > 0.05, respectively. Contrastingly, elevated pre-treatment NLR and LMR were also observed among patients with late-onset BC compared to patients with early-onset BC (p< 0.05 and p> 0.05, respectively. The pre-treatment PNLR, NLR, and TWBC were significantly higher among patients who received care for 6 months or less compared to patients who received care for more than 12 months (p< 0.05). Contrastingly, the pre-treatment LMR was insignificantly lower among patients who received care for 6 months or less compared to patients who received care for more than 12 months (p> 0.05).

### Correlation between Socio-clinical characteristics

The study found that there were direct relationships between parity and late-onset BC (r= 0.415, p= 0.000), late-stage BC and age of first pregnancy (r= 0.251, p= 0.048), and a higher level of education and employment rate (r= 0.241, p= 0.015). On the other hand, there were inverse relationships between parity and high tumour score (r= -0.319, p= 0.023), increasing level of education and parity (r= -0.420, p= 0.000), increasing level of education and late-onset BC (r= -0.368, p= 0.000), age at first pregnancy and late-onset BC (r= -0.264, p= 0.029), and employment rate and late-onset BC (r= -0.169, p= 0.042). The findings of this study revealed direct relationships between late-stage BC and high-grade tumours (r= 0.271, p= 0.028), as well as late-stage BC and in-hospital death (r= 0.396, p = 0.002). Additionally, there was a correlation between taking oral contraceptives and developing high-grade tumours (r= 0.313, p= 0.008), as well as between having an elevated TWBC before treatment and having a high body mass index (r = 0.286, p = 0.008). Furthermore, the study revealed that having an elevated neutrophil-to-lymphocyte ratio before treatment was associated with having an elevated platelet-to-lymphocyte ratio and recurrent BC (r= 0.476, p= 0.000), while there was an inverse relationship between age and the frequency of Luminal A and TNBC (r=-0.367, p= 0.030). The study shows that in-hospital death was inversely related to the number of chemotherapy courses (r= -0.310, p= 0.016), and having an elevated lymphocyte-to-monocyte ratio before treatment was inversely associated with the frequency of Luminal A and TNBC (r= -0.425, p= 0.048).

## Discussion

To identify factors associated with the high mortality rate in West Africa and identify prognostic tools, this study compared the sociodemographic and clinicopathologic features, p53 and BRCA2 expressions, hormone receptors and HER2, and systemic inflammatory indices in early-onset and late-onset BC in West Africa. This study found that the number of BC diagnosed in 2019 was twice or more than the number diagnosed in other years. The reason for the increased diagnosis is unknown. A systematic review showed that there was a decrease in cancer diagnoses during the COVID-19 pandemic [13]. Another study reported an increased mortality rate of breast cancer patients from 2019 to 2021 due to reduced screening during the COVID-19 pandemic [14]. This might be the explanation for the lower survival duration among the patients from 2020 to 2022 compared to 2017 to 2019. Additionally, the reduction in screening during the pandemic may be the reason for the higher number of early-onset BC observed from 2020 to 2022.

This study also found that patients who developed breast cancer at an earlier age had a higher level of education as compared to those who developed it later in life. This might explain the lower incidence of advanced stage III/IV breast cancer (1.4%) in the former group than the latter. The findings emphasize the impact of education on breast cancer screening uptake [15]. The noteworthy observation here is the elevated rate of unemployment among the patient population, particularly within the subset of patients with late-onset breast cancer. Furthermore, there is a notable disparity in the utilization of treatment options, specifically the completion of full courses of chemotherapy and the uptake of second-line chemotherapy, between unemployed patients and their employed counterparts. Collectively, it can be posited that the heightened unemployment rate among patients with late-onset BC may contribute to the reduced adherence to both initial and subsequent chemotherapy treatments, ultimately diminishing overall patient survival rates.

While there was no discernible distinction in alcohol consumption between patients with early-onset and late-onset breast cancer, a noteworthy contrast emerged concerning tobacco usage. Notably, the patients had ceased smoking before their cancer diagnosis, suggesting that early-life exposure to tobacco may heighten the risk of breast cancer development in later years. It’s essential to highlight that a significant 80% of the patients were overweight (≥24.9 kg/m^2^), a finding consistent with prior studies associating a personal history of smoking with the prediction of breast cancer incidence and mortality [16,17]. A high percentage of the patients in this study were obese (≥30 kg/m^2^; 35.6%), especially among patients with late-onset BC. Although the prevalence of diabetes mellitus was low, it was relatively high among patients with late-onset breast cancer. It is worth noting that obesity and diabetes are known to be strongly associated with an increased risk of developing breast cancer, as highlighted by Kang *et al*. [18]. This study also revealed that the risk of early-onset BC was higher in women who experience late menarche, delay their first pregnancy, and have low parity, compared to patients with late-onset. These risk factors are associated with prolonged exposure to estrogen, a known contributor to breast cancer development [19,20].

In this study, it is noteworthy that the prevalence of a family history of cancer exceeds the rates reported for African-American women diagnosed with breast cancer (10.3%) in the study conducted by Bethea *et al*. [21]. Similarly, the observed prevalence is higher than the 6.6% reported in Uganda and Cameroon, as noted by Adedokun *et al*. [22]. Among the subset of patients with a family history of cancer, a substantial 76.5% had first-degree relatives affected, while 23.5% had second-degree relatives with a history of cancer. Furthermore, within this group, the prevalence of early-onset breast cancer stood at 70.6%, indicating that inherited gene mutations likely contribute to an elevated risk of early-onset breast cancer. Additionally, it is noteworthy that the frequency of BRCA2 loss in this study surpasses the 5.6% prevalence recorded among patients in Uganda and Cameroon according to the findings of Adedokun *et al*. [22]. These results underscore the significance of genetic factors in breast cancer susceptibility, particularly in the context of familial cancer histories. Although the prevalence of oral contraceptive use in this study is low, literature has shown that it increases the risk of developing breast cancer [23].

In this study, a high frequency of TNBC was observed compared with other molecular subtypes of BC, especially among patients with early-onset BC. The frequency of TNBC recorded in this study (34.2%) is higher than the frequency of 24.7% and 27% reported by Akakpo *et al*. [24] and Huo *et al*. [25] in Ghana and African women, respectively but lower than the frequency (82%) reported by Stark *et al*. [26] among Ghanaians. The reason for the difference is unknown but could be associated with the mutation of DNA repair genes such as BRCA2 and p53 or social characteristics. Additionally, the frequency of TNBC and Luminal A were higher among patients with early-onset BC compared with late-onset BC. This agrees with the findings of Akakpo *et al*. [24] in Ghana. In this study, the highest frequency of obesity was found among patients with Luminal A breast cancer (85.7%). In contrast, Vona-Davis et al. [27] observed higher cases of obesity among patients with TNBC (50%) than non-TNBC cases (16%).

The frequency of p53 expression in TNBC cases is like the prevalence rate reported in Ghana (31.0%) by Ameh-Mensah *et al*. [28] but lower than the frequency rate reported in Morocco (41.0%) by Jouli *et al*. [29]. Here, the frequency of BRCA2 loss in TNBC is higher than the prevalence rate reported by Jiagge *et al*. [30] and Xie *et al*. [31] in Ghana (40%) and China (20%), respectively. The difference in BRCA2 expression in TNBC could be race and lifestyle-related. Studies have shown that a high expression of the p53 gene is associated with high-grade large-size and highly proliferative tumours with lymph node involvement, and risk of recurrence [28,29,32]. Thus, the concomitant high expression of p53 and moderate to low expression of BRCA2 in this study could be an early immune response to aggressive tumours. According to Lee et al., individuals with loss of both BRCA2 and p53 function are at risk of early-onset BC [33]. The latter might be an explanation for the lower mean age of TNBCs compared with Luminal B-like cancer. The absence or low expression of p53 and BRCA2 in a malignant breast tumour suggests poor prognosis.

This study revealed elevated pre-treatment PLR among patients with early-onset BC compared with late-onset BC. This study also found that elevated pre-treatment NLR, PNLR and TWBC were associated with shorter survival rates. Cho et al. revealed that the PLR, an indicator of systemic inflammation as a part of the host immune response, was an independent marker for poor disease-free survival in patients with lymph node metastasis and luminal subtype [34]. Finally, our study is limited by a small sample size and single-centre origin. Future studies should collect data from multiple tertiary healthcare facilities and a larger patient sample.

## Conclusion

This study provides valuable insights into factors affecting BC diagnosis, prognosis, and treatment outcomes in West Africa, shedding light on the complex interplay of sociodemographic, clinical, genetic, and immunological factors in BC patients. This study found a high incidence of early-onset BC, p53 mutation, and TNBC. Additionally, it suggests that pre-treatment systemic inflammatory indices can identify high-mortality-risk patients and early-onset BC. These findings can inform strategies for early detection, treatment, and support for individuals facing BC in the region. It suggests that TWBC, PNLR, and NLR should be used to monitor patients to improve treatment outcomes.

## Data Availability

All data produced in the present study are available upon reasonable request to the authors

## Acknowledgements

Special thanks are to the Histopathology Unit, Medical Laboratory Services, University of Benin University Teaching Hospital and Nnamdi Azikiwe University Teaching Hospital staff for their technical assistance.

## Data availability

Data will be made available by the corresponding author upon reasonable request.

## Ethics Statement

The protocol of this retrospective study was approved by the Institutional Review Board (NAUTH/CS/66/VOL.14/VER.3/159/2021/124 and NAUTH/CS/66/VOL.14/VER.3/106/ 2021/109). The study was conducted according to the Declaration of Helsinki by the World Medical Association (WMA) General Assembly.

